# OHCA-EXTRACT: Evaluating the Accuracy of a Large Language Model Pipeline for Out-of-Hospital Cardiac Arrest Case Identification and Utstein Variable Extraction

**DOI:** 10.64898/2026.06.29.26356890

**Authors:** Aditya C Shekhar, Donald Apakama, Aryan Saharan, Kevin Petrozzo, Joy Jiang, Alexis Zebrowski, David Buckler, Ryan Huebinger, Benjamin S Abella, Lynne D Richardson, Girish N Nadkarni, Ethan E Abbott

**Affiliations:** Icahn School of Medicine at Mount Sinai, New York, NY; Department of Emergency Medicine, Icahn School of Medicine at Mount Sinai, New York, NY; Windreich Department of Artificial Intelligence and Human Health, Icahn School of Medicine at Mount Sinai, New York, New York, USA; Charles Bronfman Institute for Personalized Medicine, Icahn School of Medicine at Mount Sinai, New York, New York, USA; University of New Mexico Health Science Center, Albuquerque, New Mexico, USA; The Hasso Plattner for Digital Health at Mount Sinai Icahn School of Medicine at Mount Sinai, New York, New York, USA; Department of Population Health Science and Policy, Icahn School of Medicine at Mount Sinai, New York, NY; Institute for Health Equity Research (IHER), Icahn School of Medicine at Mount Sinai, New York, NY

**Keywords:** Out-of-hospital cardiac arrest, Utstein template, large language models, clinical informatics, automated data extraction, artificial intelligence

## Abstract

**Background:** Manual identification and abstraction of out-of-hospital cardiac arrest (OHCA) cases and Utstein template variables from electronic health records is resource-intensive and limits scalable measurement for observational research and registry participation. We evaluated a large language model (LLM)–assisted pipeline to identify true OHCA encounters within an ICD-coded emergency department (ED) cohort and to also extract a limited set of Utstein variables from unstructured documentation, with iterative pipeline refinement and final performance evaluation conducted on the same validation cohort.

**Methods:** We conducted a study of ICD-flagged OHCA encounters within a large urban academic health system (2015–2024). A two-module pipeline was developed that included an identification module and a variable-extraction module. The pipeline was evaluated against independent manual chart abstraction on a validation sample (n = 176) with physician adjudication. The identification module performed binary OHCA classification along with etiology classification. The variable-extraction module extracted five Utstein variables: witnessed status, EMS defibrillation, first recorded rhythm, arrest location, and bystander response. We calculated sensitivity, specificity, PPV, NPV, and F1 scores (Clopper–Pearson 95% CIs) using the manual chart abstraction as the reference gold standard.

**Results:** Of 176 processed encounters, 152 had complete gold-standard classification and were included in identification analyses (OHCA prevalence 81.6%; n = 124 true OHCA, n = 28 non-OHCA). The identification module achieved an accuracy of 0.91 (95% CI 0.85–0.95), sensitivity 0.94 (0.89–0.98), specificity 0.75 (0.55–0.89), PPV 0.94 (0.89–0.98), NPV 0.75 (0.55–0.89), and F1 0.94, compared with accuracy 0.82 for a prevalence-only baseline. Variable-level accuracy ranged from 0.63 to 0.75 (Kappa range 0.31–0.51) across the five extracted Utstein variables, with bystander response showing the lowest agreement (accuracy 0.63, Kappa = 0.31).

**Conclusions:** Within-sample performance estimates indicate that this LLM pipeline can identify true OHCA encounters within an ICD-flagged ED cohort with accuracy and PPV above an administrative-coding baseline. Variable-level extraction accuracy ranged from fair to moderate across the five evaluated Utstein variables, indicating that further methodological development is required before automated extraction is feasible. With external validation and integration with targeted human verification, this approach may support human-augmented workflows using LLMs, however independent external validation is required before our findings can be generalized.

## Introduction

Out-of-hospital cardiac arrest remains one of the most critical time-sensitive emergencies, affecting more than 400,000 individuals annually in the United States.^1,2^ Despite advances in prehospital care and post-resuscitation management, survival to hospital discharge and survival with good neurological outcomes remain low.^2–4^ Improving outcomes requires accurate, standardized data collection to identify modifiable factors in the resuscitation chain of survival and enable comparison of system performance across regions and institutions. Further, current US registry datasets lack the ability to connect granular clinical data across pre-hospital, hospital, and post-discharge to establish long-term and longitudinal outcomes.^5,6^

The Utstein template, established in 1991 and endorsed by the American Heart Association (AHA), European Resuscitation Council, and International Liaison Committee on Resuscitation (ILCOR), provides a standardized framework for OHCA data collection. Core variables encompass prehospital variables such as witnessed status, bystander cardiopulmonary resuscitation (CPR), initial cardiac rhythm, and time intervals. These standardized fields also include resuscitation interventions, subsequent in-hospital care, and patient outcomes, forming the backbone of cardiac arrest registries (e.g., the Cardiac Arrest Registry to Enhance Survival [CARES] in the United States).^7,8^

While manual abstraction remains the mainstay of OHCA registry data collection, each case may require a significant number of resources and abstraction time, making the process expensive, potentially error-prone, and challenging to scale.^9,10^ Consequently, many health systems and hospitals, particularly smaller community hospitals and rural health systems, cannot justify contributing data to cardiac arrest registries, leaving significant gaps in OHCA data collection that affect both research and quality improvement.^7,9^ Traditional rule-based natural language processing (NLP) systems have shown some promise for clinical variable extraction from unstructured clinical documentation but require substantial customization and struggle with temporal reasoning and contextual ambiguity.^11,12^ In contrast, large language models (LLMs) demonstrate strong contextual reasoning and can generalize across diverse documentation styles without extensive retraining or pre-processing.^9,12,13^ However, LLM approaches compared with manual abstraction have not been rigorously quantified in real-world health system deployments, especially for OHCA-specific data.^12,14^

This study presents OHCA-EXTRACT, a dual-module LLM pipeline designed to automate identification and Utstein-variable extraction for OHCA encounters from a large, multicenter academic health system in a major metropolitan area. The objective was to evaluate the accuracy of the pipeline to (1) identify true OHCA encounters from unstructured electronic health record (EHR) documentation within an ICD-flagged ED cohort, and (2) extract a subsample of key Utstein variables, validating LLM performance against a gold-standard human-abstracted reference.

## Methods

### Study setting, cohort, and data sources

The Mount Sinai Health System is a large, integrated academic medical center in New York City serving a diverse urban population. We retrospectively examined data for clinical encounters between January 1, 2015, and December 31, 2024. The candidate cohort was first identified using a structured query language (SQL) query to flag ED encounters with a primary or secondary diagnosis containing cardiac arrest–related ICD-10 codes: I46.8, I49.01, R00.1, T82.199A, T75.4XXA, I97.120, I46.2, I46.9, and I97.121.^15,16^ When a patient had multiple qualifying encounters across the study period, we used the earliest qualifying (index) OHCA encounter associated with documented arrest as the primary unit of analysis. For each index encounter, all available unstructured clinical documentation was retrieved and aggregated at the encounter level. Documentation sources included EMS/prehospital narratives, ED triage documentation, ED clinician notes, nursing notes, code/event records, inpatient notes, and disposition documentation. Structured tabular data included age, sex, gender, race and ethnicity, and comorbidities at the time of arrest.

### Reference standard manual chart abstraction

To validate our results, we first created a human-annotated reference standard dataset to test against the LLM pipeline. We identified a random sample of 200 OHCA patient-level encounters within the EHR data for manual abstraction. No formal a priori sample size calculation was performed, and the 200-encounter manual abstraction sample was selected to balance annotation resource constraints with the goal of characterizing pipeline performance across a clinically representative range of OHCA encounters. After exclusions for missing key documentation or inaccessible records, we included a validation sample of 176 OHCA encounters for human annotation. This validation cohort served as the gold-standard reference for LLM performance assessment. Three clinical research abstractors who received formal training in OHCA chart abstraction methodology independently reviewed each patient’s clinical documentation and abstracted the data into a Research Electronic Data Capture (REDCap) instrument. A data dictionary was developed specifically for this research and leveraged Utstein template specifications, encompassing more than 70 Utstein-aligned variables organized into five primary data entry forms: demographics and baseline characteristics (age, sex, race/ethnicity, comorbidities), prehospital Utstein variables (witnessed status, arrest location, bystander CPR, first monitored rhythm, etiology classification, time intervals), in-hospital clinical data (EMS interventions, hospital procedures, treatments, consultations), withdrawal of life-sustaining treatment data, and discharge disposition and neurological outcomes. A standardized abstraction manual provided detailed definitions for each variable, coding rules, and decision trees for ambiguous scenarios, all aligned with official Utstein template specifications and clinical guidelines. *For this initial pipeline evaluation, we selected five core Utstein fields including witnessed status, EMS defibrillation, first recorded rhythm, arrest location, and bystander response.* These were chosen as they represent core Utstein variables and provide a representative mix of binary, ternary, and categorical structures. Overall percent agreement was calculated as the ratio of agreed variable-level codings to total codings across these five Utstein fields used for LLM evaluation. Fleiss’ kappa across the three abstractors was computed and interpreted using Landis–Koch thresholds. Discrepancies between abstractors were adjudicated by an emergency medicine physician (EA), blinded to LLM output, to establish the final reference standard.

### Large Language Model Pipeline Architecture

We engineered a two-step LLM pipeline that included two modules: (1) an identification module to classify each encounter as true OHCA versus non-OHCA, and (2) a variable-extraction module to extract the subset of prespecified Utstein variables. The pipeline was implemented using an institution-specific secure Azure instance and accessed OpenAI GPT-4o-mini (model version 2024-07-18; Azure OpenAI API version 2024-08-01-preview) via API. GPT-4o-mini was selected to balance per-encounter cost, latency, and throughput for a pipeline intended to scale to tens of thousands of encounters. A formal head-to-head comparison against larger frontier models was not performed in this evaluation. The focus of this work was operationalizing the pipeline with future efforts aimed at benchmarking different models. Use of a HIPAA-compliant Azure instance allowed for processing of complete clinical documentation with minimal de-identification. All model calls used deterministic settings (temperature = 0) and were constrained to structured JSON outputs validated against Pydantic schemas. Records failing schema validation were flagged and excluded from analysis

For pipeline development, a context engineering approach was used for inputs. This included all structured variable definitions and coding rules from the abstractor data dictionary, aligning the model’s reasoning framework with that of trained human abstractors. Clinical note text was preprocessed with section filtering and token-budget truncation prior to submission. The model was not exposed to the full corpus of available clinical documentation for a given encounter. Inputs were limited to a curated subset selected by note type, further filtered at the section level, and truncated to a maximum of 1,800 tokens per encounter. The 1,800-token cap was set empirically during pipeline development to balance cost against the risk of excluding relevant OHCA narratives.

### Identification Module

The identification module returned a structured JSON record containing three fields: (1) a binary OHCA classification (ohca_yes_no), (2) an etiology code drawn from an eight-category enumeration (1 = cardiac [presumed cardiac cause]; 2 = traumatic; 3 = overdose; 4 = other medical; 5 = in-ED arrest; 6 = inpatient arrest; 7 = unknown etiology; 8 = unknown location), and (3) a continuous confidence score (range 0–1) representing the model’s self-reported certainty in its classification. The enumeration is intentionally hybrid across etiology and location given this data is often captured together in unstructured narratives. Codes 1–4 and 7 capture clinical etiology, codes 5 and 6 capture arrest location after ED arrival, and code 8 applies when neither etiology nor location can be determined from the documentation. Codes 5 and 6 function as explicit OHCA disqualifiers and force ohca_yes_no = 0, serving as the mechanism by which the module distinguishes true OHCA from emergency department and inpatient arrests with overlapping or incorrect ICD codes. Codes 1–4, 7, and 8 do not force a particular ohca_yes_no value, and the binary flag is assigned independently when these codes are used. This hybrid design reflects the operational use of location as a disqualifier, which is the mechanism by which the pipeline gains specificity over administrative coding alone. A prespecified fallback automatically resubmitted any encounter with a confidence score below 0.7 using an expanded 3,000-token window. No encounter met the <0.7 threshold, and the fallback did not fire (0 of 176). The distribution of confidence scores and their relationship to classification accuracy were evaluated as a secondary outcome. In this initial evaluation, performance was formally assessed only for ohca_yes_no. (full system prompt and JSON schema for the identification module are provided in Supplement S1)

### Variable Extraction Module

The variable-extraction module used two separate prompts per encounter. A primary extraction call returned witnessed status, arrest location, EMS defibrillation, and first recorded rhythm as a single Pydantic-validated JSON object. A separate two-stage standalone prompt was used for bystander CPR: stage 1 returned verbatim CPR-related quotes from the clinical note, and stage 2 coded the variable from those quotes alone, with no access to the surrounding text. The bystander prompt was separated from the primary call to allow stricter attribution rules. Within the primary call, witnessed status was extracted single-pass, while EMS defibrillation, first recorded rhythm, and arrest location used quote-grounded extraction in which the schema required the model to return supporting verbatim text from the clinical note alongside each classification. When no supporting text could be identified, the model was instructed to return an explicit “not found” value in the quote field rather than a classification. Such records were treated as missing for that variable in the both-present analysis. Witnessed status was extracted single-pass because pilot testing showed quote-grounding did not improve agreement for this binary variable. The primary extractor produces an Utstein-aligned record. The five fields described below were chosen for this initial pipeline evaluation as core Utstein variables representing a mix of binary, ternary, and categorical types. Performance on the remaining fields was not assessed in this analysis.

Witnessed status was classified as binary (witnessed vs. not witnessed by non-EMS person). First recorded rhythm was classified as a categorical variable into nine categories, including ventricular fibrillation, pulseless ventricular tachycardia, ventricular tachycardia with pulse, pulseless electrical activity, asystole, and bradycardia, plus unknown shockable, unknown nonshockable, and unknown/not recorded subcategories. EMS defibrillation was a ternary variable (yes / no / unknown) recording whether defibrillation was delivered by EMS in the field. Arrest location was a categorical variable capturing setting of arrest (home/residence, public location, healthcare facility, other).

Bystander response was a ternary variable (yes / no / unknown) recording whether chest compressions were performed by a non-EMS person (family, layperson, police, security, or dispatcher-assisted caller) prior to EMS arrival. Compression-only and conventional bystander CPR both counted as “yes.” Bystander AED use without compressions was captured under a separate variable and was not evaluated here. The model was instructed to assign “yes” only when pre-EMS compressions by a non-EMS person were explicitly documented, “no” when documentation stated CPR was initiated by EMS on arrival, and “unknown” when attribution or timing was ambiguous. (System prompts and schemas for both the primary extraction call and the standalone two-stage bystander CPR call are provided in Supplement S2.)

### Context engineering

The pipeline was developed iteratively, with context engineering serving as the primary mechanism for aligning model behavior to the abstractor reference standard. Context engineering is an approach similar to model prompting that involves specifying the inputs, including which note types are leveraged, section filtering, the structured variable definitions and coding rules injected into the prompt, and the output schema constraints. Rather than fine-tuning model weights, we refined this input frame across successive pipeline runs on encounters drawn from the same cohort used for final evaluation. Refinement was driven by inspection of misclassified encounters after each run. Two categories of systematic error recurred early in development and shaped the final prompt and schema. First, in-hospital cardiac arrests — including both arrests occurring after ED triage and inpatient arrests on hospitalized patients — were initially classified as OHCA by the model when ICD-coded cardiac arrest diagnoses appeared in the chart without clear prehospital context. This pattern motivated the addition of etiology codes 5 (in-ED arrest) and 6 (inpatient arrest) to the identification schema, with an explicit post-hoc rule to identify true OHCA cases with high accuracy. Second, bystander CPR attribution was inconsistent when notes described chest compressions without identifying whether they were performed by EMS, which initially led to misclassification. Each change was implemented in the prompt, schema, or post-processing layer without adjusting model weights, and was retained when it reduced misclassification without introducing offsetting errors elsewhere. We did not hold out a separate development set prior to the validation run and iterative refinement and final evaluation were conducted on the same cohort. The reported performance estimates should therefore be interpreted with caution as this approach did not include external validation.

### Model Evaluation and Statistical Analysis

For the identification module, we calculated accuracy, sensitivity, specificity, positive predictive value, negative predictive value, and F1 scores, with 95% confidence intervals. As a reference comparator we constructed a prevalence-only baseline naïve classifier that assigns ‘ohca_yes_no = 1’ to every ICD encounter without reading clinical documentation, representing the performance achievable by accepting administrative coding alone. By construction this baseline has sensitivity 1.0 and undefined specificity, with accuracy equal to cohort OHCA prevalence. Differences between the identification module and this baseline are reported descriptively as absolute differences in percentage points. No formal hypothesis testing was performed for these comparisons. For the variable-extraction module, we report variable-level accuracy and Cohen’s Kappa using a both-present analysis restricted to encounters in which both the LLM and the human abstractor returned a non-missing value. As a prespecified sensitivity analysis, we additionally recomputed accuracy treating any model-missing value as incorrect over all gold-available encounters and report per-variable model completeness (coverage). Confidence-score distributions were summarized descriptively and examined for association with classification error. All analyses were performed in Python 3.13.

### Study Approvals

The study was reviewed and approved by the Institutional Review Board at the Icahn School of Medicine at Mount Sinai (STUDY-24-01201). Informed consent was not required given the retrospective nature of this study. This study was reported in accordance with the Transparent Reporting of a Multivariable Prediction Model for Individual Prognosis or Diagnosis with Artificial Intelligence (TRIPOD+AI) guideline.^17^ A completed checklist is provided as Supplement S4.

## Results

### Reference Dataset and Abstractor Agreement

Of 176 validation encounters with manual abstraction, 152 had complete gold-standard OHCA classification and were included in classifier performance analyses (Figure 1). The remaining 24 were excluded for missing or incomplete data needed for comparison. Across the three abstractors, overall variable-level agreement was 88.4% and Fleiss’ Kappa was 0.69, corresponding to substantial agreement (Figure 2).

**Figure 1.**
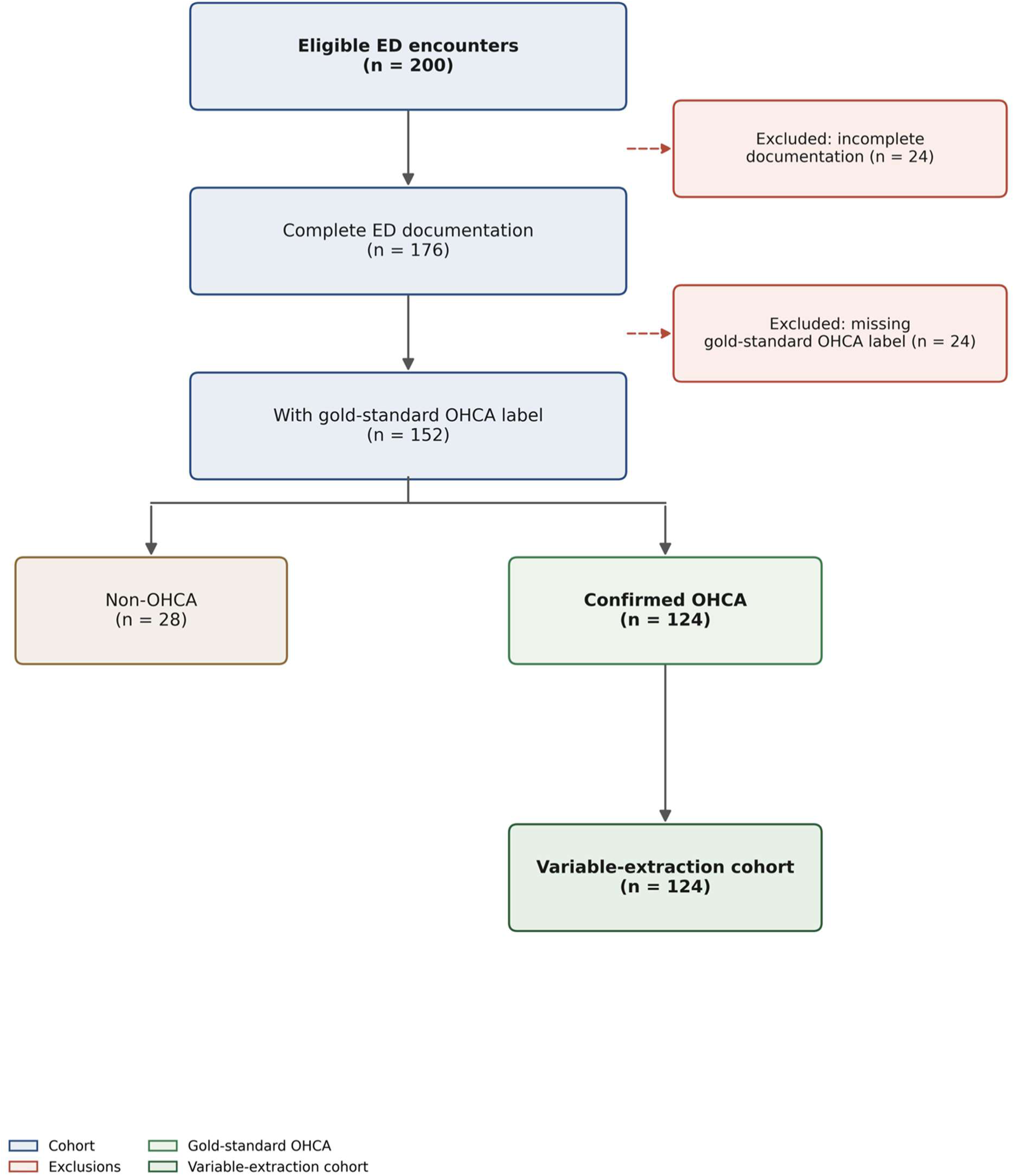
Study flow diagram. Figure 1 Legend: Flow of encounters from initial cohort identification through final analytic samples. A random sample of 200 OHCA patient-level encounters was drawn from the ICD-flagged candidate cohort (Mount Sinai Health System, 2015–2024) for manual abstraction. After exclusions for missing key documentation, 176 encounters comprised the validation sample for human annotation. Of these, 152 had complete gold-standard OHCA classification and were included in identification-module performance analyses (n = 124 confirmed OHCA, n = 28 non-OHCA); 24 were excluded for missing or incomplete data needed for comparison. The 124 confirmed OHCA encounters were used for variable-extraction analyses and were all evaluable at the variable level (no schema-validation failures). Abbreviations: OHCA, out-of-hospital cardiac arrest; ICD, International Classification of Diseases.

**Figure 2.**
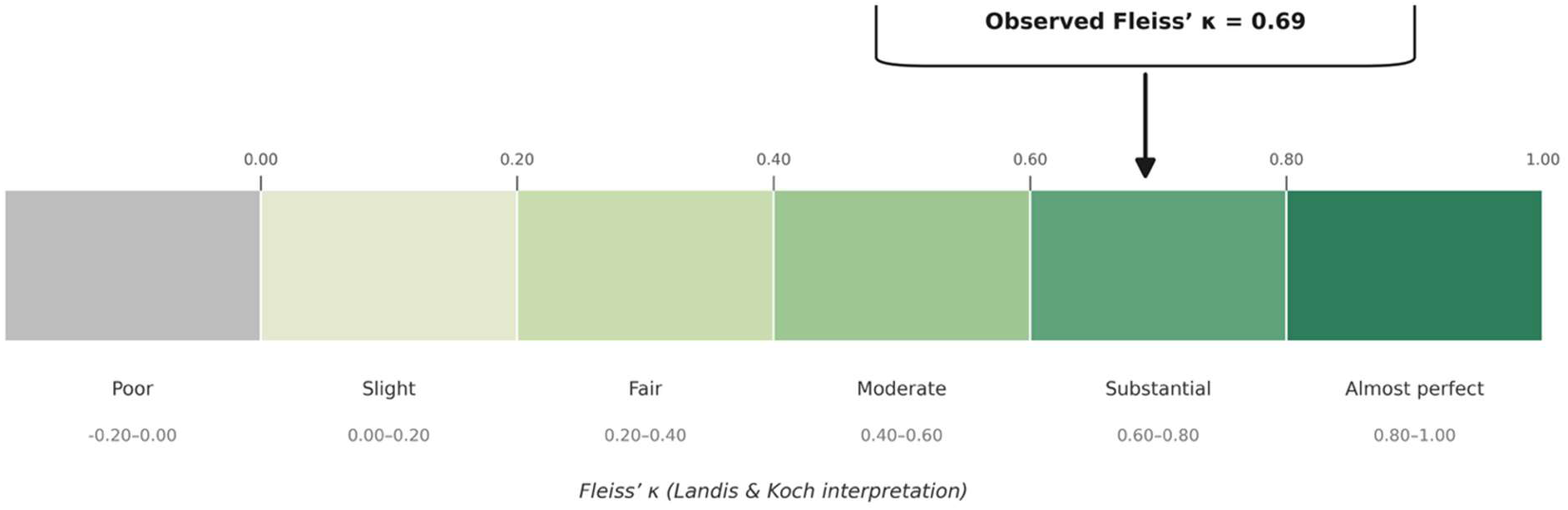
Reference-standard inter-rater reliability. Figure 2 Legend: Inter-rater reliability among three independent clinical research abstractors who reviewed the 176-encounter validation cohort. Overall variable-level percent agreement was 88.4%. Fleiss’ Kappa across the three abstractors was 0.69, corresponding to substantial agreement on the Landis–Koch scale (κ ≥ 0.60). Discrepancies were adjudicated by an emergency medicine physician blinded to LLM output to establish the final gold-standard reference.

### Identification Module performance

Among the 152 analyzed encounters, OHCA prevalence was 81.6% (n = 124 true OHCA; n = 28 non-OHCA). The identification module (Figure 3) correctly classified 138 of 152 encounters, yielding accuracy 0.91 (95% CI 0.85–0.95) (Table 1). Of 124 confirmed OHCA cases, 117 were identified (sensitivity 0.94, 95% CI 0.89–0.98), with 7 false negatives. Of 28 non-OHCA cases, 21 were correctly ruled out (specificity 0.75, 95% CI 0.55–0.89), with 7 false positives. PPV was 0.94 (95% CI 0.89–0.98) and NPV was 0.75 (95% CI 0.55–0.89). F1 was 0.94. Relative to the prevalence-only baseline (accuracy 0.82, F1 0.90, specificity undefined), the identification module improved accuracy by 9.2 percentage points, PPV by 12.8 percentage points, and F1 by 4.5 percentage points, while recovering a specificity estimate that administrative coding alone cannot produce.

**Figure 3.**
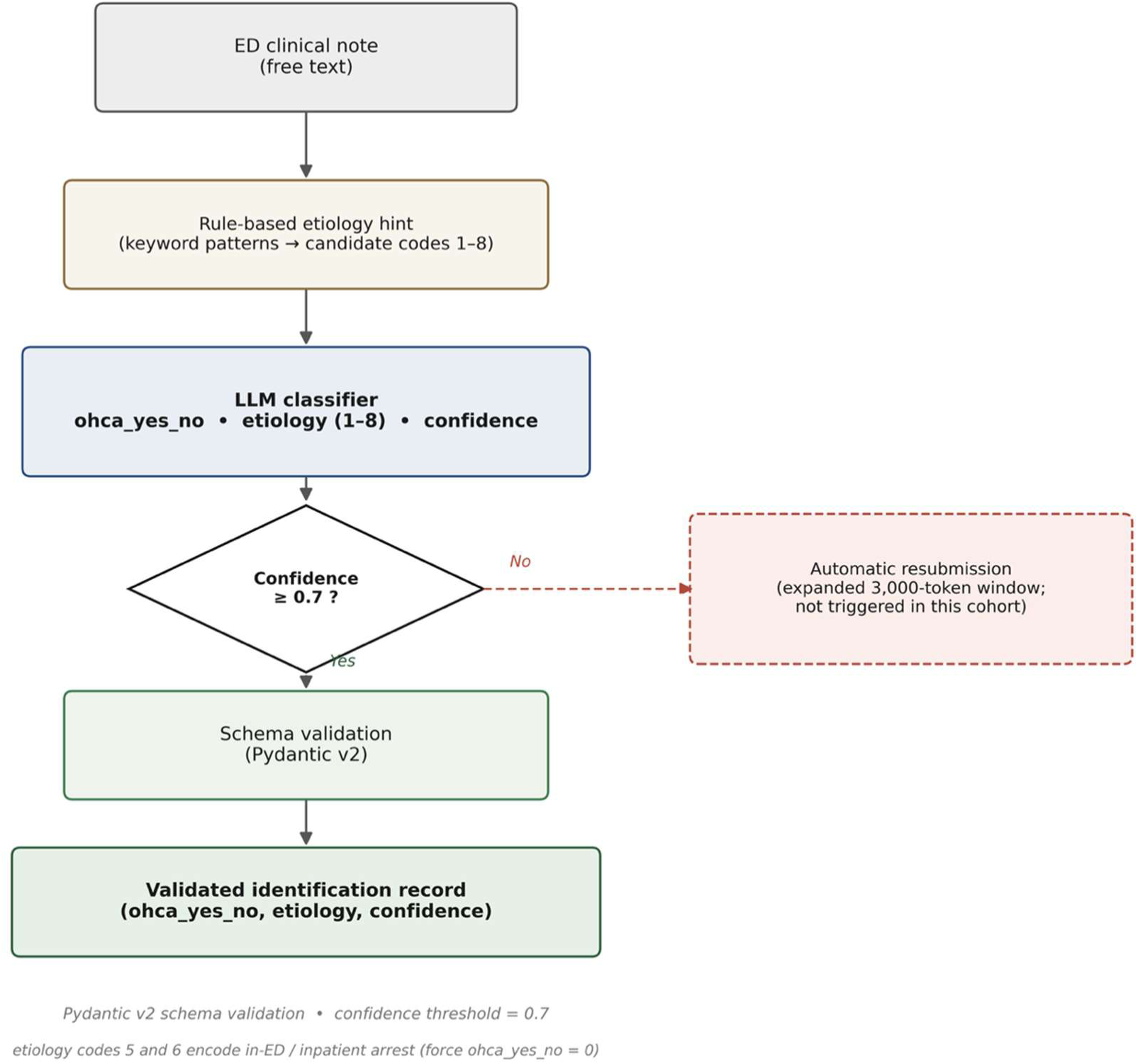
Identification module pipeline. Figure 3 Legend: Schematic of the OHCA identification module. ICD-flagged ED encounters enter the pipeline; clinical documentation is preprocessed via note-type selection, section filtering, and truncation to a maximum of 1,800 tokens. The preprocessed text is passed with structured variable definitions and coding rules to the GPT-4o-mini model (temperature = 0; structured JSON output). The module returns three fields: a binary OHCA classification (‘ohca_yes_nò), an eight-category etiology code, and a continuous self-reported confidence score (0–1). Etiology codes 5 (in-ED arrest) and 6 (inpatient arrest) force ‘ohca_yes_no = 0’, encoding arrest location as the mechanism by which the module distinguishes true OHCA from in-hospital arrests with overlapping ICD codes. Encounters with confidence < 0.7 were eligible for automatic resubmission with an expanded 3,000-token window. Abbreviations: OHCA, out-of-hospital cardiac arrest; ED, emergency department; ICD, International Classification of Diseases; JSON, JavaScript Object Notation.

**Table 1.** Identification Module — Case-Level OHCA Classification Performance. Table 1 Legend: Case-level classification performance of the LLM identification module against the manually adjudicated gold-standard reference (n = 152 encounters with complete gold-standard OHCA classification). Confusion matrix: true positives = 117; false negatives = 7; false positives = 7; true negatives = 21. Of 176 total processed encounters, 24 were excluded for missing gold-standard OHCA classification; no encounters were excluded for LLM processing errors. Performance metrics are reported with 95% Clopper–Pearson exact binomial confidence intervals. Prevalence-only baseline: a naïve classifier that assigns OHCA = 1 to every ICD-flagged encounter, representing the performance achievable by accepting administrative coding alone. By construction, the baseline has sensitivity 1.00 and undefined specificity, with accuracy equal to cohort OHCA prevalence (0.82). The Change column reports the absolute difference between the LLM classifier and the prevalence-only baseline in percentage points (pp). OHCA, out-of-hospital cardiac arrest; LLM, large language model; CI, confidence interval; PPV, positive predictive value; NPV, negative predictive value; pp, percentage points.

| Metric | LLM Classifier | 95% CI (Clopper–Pearson) | Prevalence-only baseline | Change |
| --- | --- | --- | --- | --- |
| Accuracy | 0.91 | 0.85–0.95 | 0.82 | +9.2 pp |
| Sensitivity | 0.94 | 0.89–0.98 | 1.00 | –5.6 pp |
| Specificity | 0.75 | 0.55–0.89 | — | — |
| PPV | 0.94 | 0.89–0.98 | 0.82 | +12.8 pp |
| NPV | 0.75 | 0.55–0.89 | — | — |
| F1 | 0.94 | — | 0.90 | +4.5 pp |

Examining the basis for each classification, the module’s specificity was driven almost entirely by the in-ED/inpatient location codes. The model assigned etiology code 5 (in-ED arrest) or 6 (inpatient arrest) to 27 of the 152 encounters, classifying all as non-OHCA. These captured 20 of the 21 true-negative encounters, with the remaining true negative assigned code 8 (unknown location). Treated as a discrete sub-task, code-5/6 assignment had a positive predictive value of 20/27 (0.74) for true non-OHCA. The same mechanism produced all seven false negatives. Seven gold-standard OHCA encounters were misassigned to code 5 (n = 4) or code 6 (n = 3) and thereby classified as non-OHCA. These were predominantly arrests at skilled-nursing facilities or EMS-transported arrests that the model coded as in-hospital events. No false negative was attributable to narrative truncation and in each case the model’s rationale addressed the documented arrest location, confirming the relevant text was within the input window.

### Confidence score distribution

Confidence scores were available for all 176 processed encounters and were near the ceiling of 1, with values of 0.9 (n = 72, 47.4%) or 1.0 (n = 80, 52.6%) and no score below 0.9. All 14 classification errors (7 false negatives, 7 false positives) occurred at high confidence (0.9 or 1.0). The prespecified fallback mechanism, designed to resubmit encounters with confidence below 0.7 using an expanded 3,000-token context window, did not activate for any encounter (0 of 176). The absence of any score below 0.9 and the uniform distribution of errors across the 0.9–1.0 range indicate that the confidence field had no discriminative value for identifying misclassified encounters in this evaluation. This field reflects the model’s self-reported certainty within the structured JSON output and should not be interpreted as a calibrated probability.

### Variable Extraction Module performance

The variable-extraction module (Figure 4) was applied to the 124 confirmed OHCA encounters. All were evaluable, with no schema-validation failures in the final extraction run. Performance was evaluated using a both-present analysis restricted to encounters in which both the LLM output and human abstraction provided a non-missing value (Table 2). Witnessed status showed the highest accuracy (0.75; κ = 0.47; n = 118), followed by EMS defibrillation (0.69; κ = 0.45; n = 114), arrest location (0.66; κ = 0.51; n = 117), and first recorded rhythm (0.64; κ = 0.51; n = 111). Bystander response showed the lowest agreement (0.63; κ = 0.31; n = 118) (Figure 5). Model completeness was high across variables (coverage 94–100%); in the prespecified sensitivity analysis treating model-missing values as incorrect, accuracy declined by ≤ 0.04 for every variable (Supplement S3), indicating that the both-present estimates were not materially inflated by selective omission of difficult encounters.

**Figure 4.**
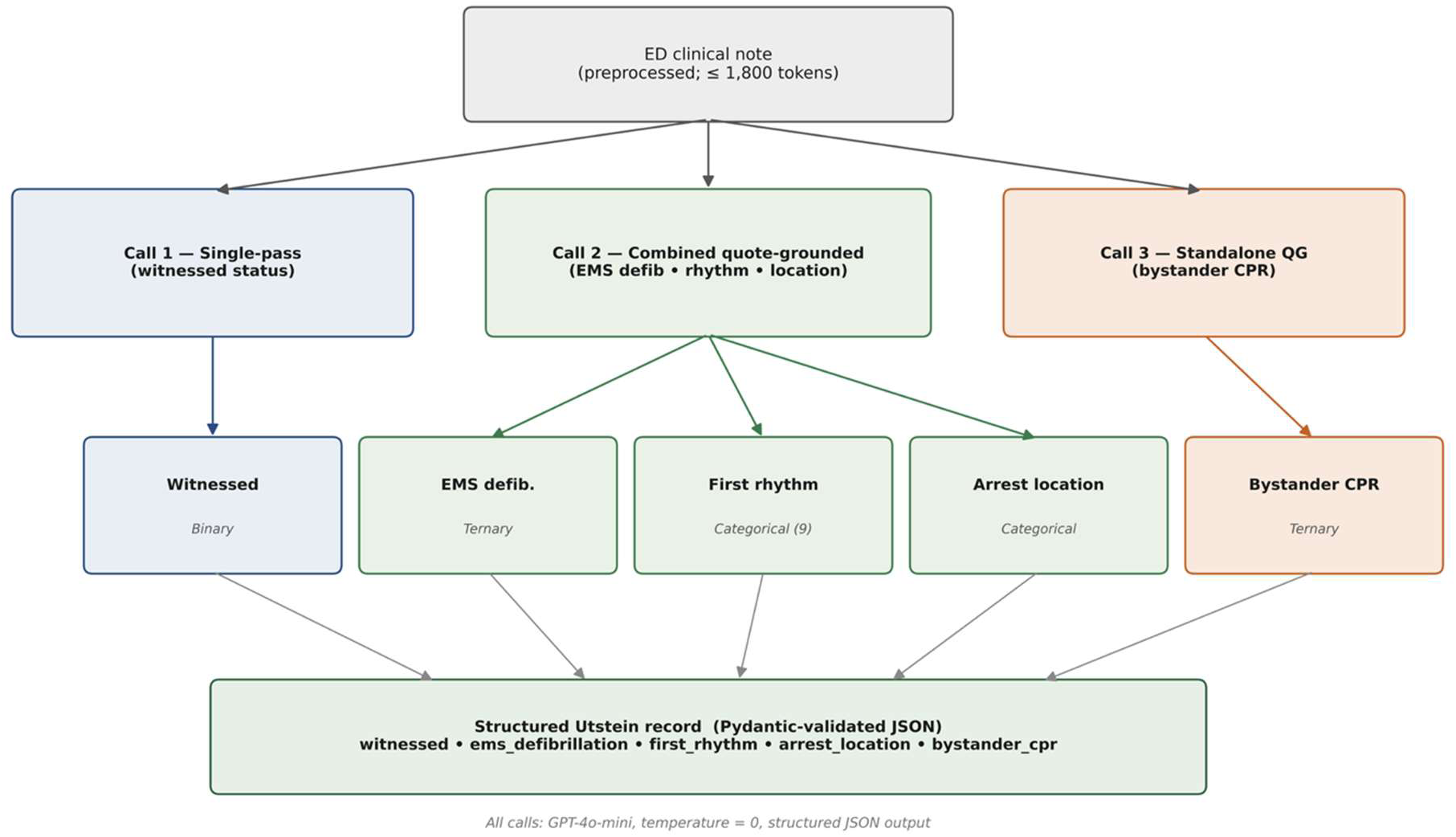
Variable-extraction module pipeline. Figure 4 Legend: Schematic of the variable-extraction module applied to encounters classified as true OHCA by the identification module. Preprocessed clinical documentation is submitted with structured field definitions, coding rules, and output schema constraints to GPT-4o-mini. The module returns a single Pydantic-validated JSON object containing five Utstein variables: witnessed status (binary), first recorded rhythm (categorical, 9 categories), EMS defibrillation (ternary), arrest location (categorical), and bystander CPR (ternary). Variables were extracted using a hybrid strategy: a single-pass call for witnessed status, a quote-grounded call requiring supporting textual evidence for first recorded rhythm, EMS defibrillation, and arrest location, and a standalone quote-grounded call for bystander CPR to allow stricter attribution rules. Abbreviations: OHCA, out-of-hospital cardiac arrest; EMS, emergency medical services; CPR, cardiopulmonary resuscitation; JSON, JavaScript Object Notation.

**Table 2.** Variable-Extraction Module: Utstein Variable Performance. Table 2 Legend: Variable-level performance of the LLM variable-extraction module against the manually adjudicated gold-standard reference, restricted to encounters classified as true OHCA (n = 124 confirmed OHCA encounters; all evaluable, with no schema-validation failures in the final extraction run). Performance is reported using a both-present analysis restricted to encounters in which both the LLM output and human abstractor provided a non-missing value; n (both present) varies by variable based on data availability. Accuracy is the proportion of encounters with matching LLM and human-abstracted values. Cohen’s Kappa is reported as a chance-corrected measure of agreement. A hybrid extraction approach was used: witnessed status was extracted using a single-pass strategy in which the model returned a direct classification; EMS defibrillation, first recorded rhythm, and arrest location were extracted using a quote-grounded strategy in which the model was constrained to return supporting textual evidence with each classification; bystander CPR was extracted using a standalone quote-grounded call, separated from the other variables to allow stricter attribution rules. Abbreviations: OHCA, out-of-hospital cardiac arrest; LLM, large language model; EMS, emergency medical services; CPR, cardiopulmonary resuscitation.

| Variable | Extraction approach | Type | n (both present) | Accuracy | Kappa |
| --- | --- | --- | --- | --- | --- |
| Witnessed | Single-pass | Binary | 118 | 0.75 | 0.47 |
| EMS Defibrillation | Quote-grounded | Ternary | 114 | 0.69 | 0.45 |
| Initial Rhythm | Quote-grounded | Categorical | 111 | 0.64 | 0.51 |
| Location | Quote-grounded | Categorical | 117 | 0.66 | 0.51 |
| Bystander CPR | Quote-grounded (standalone) | Ternary | 118 | 0.63 | 0.31 |

**Figure 5.**
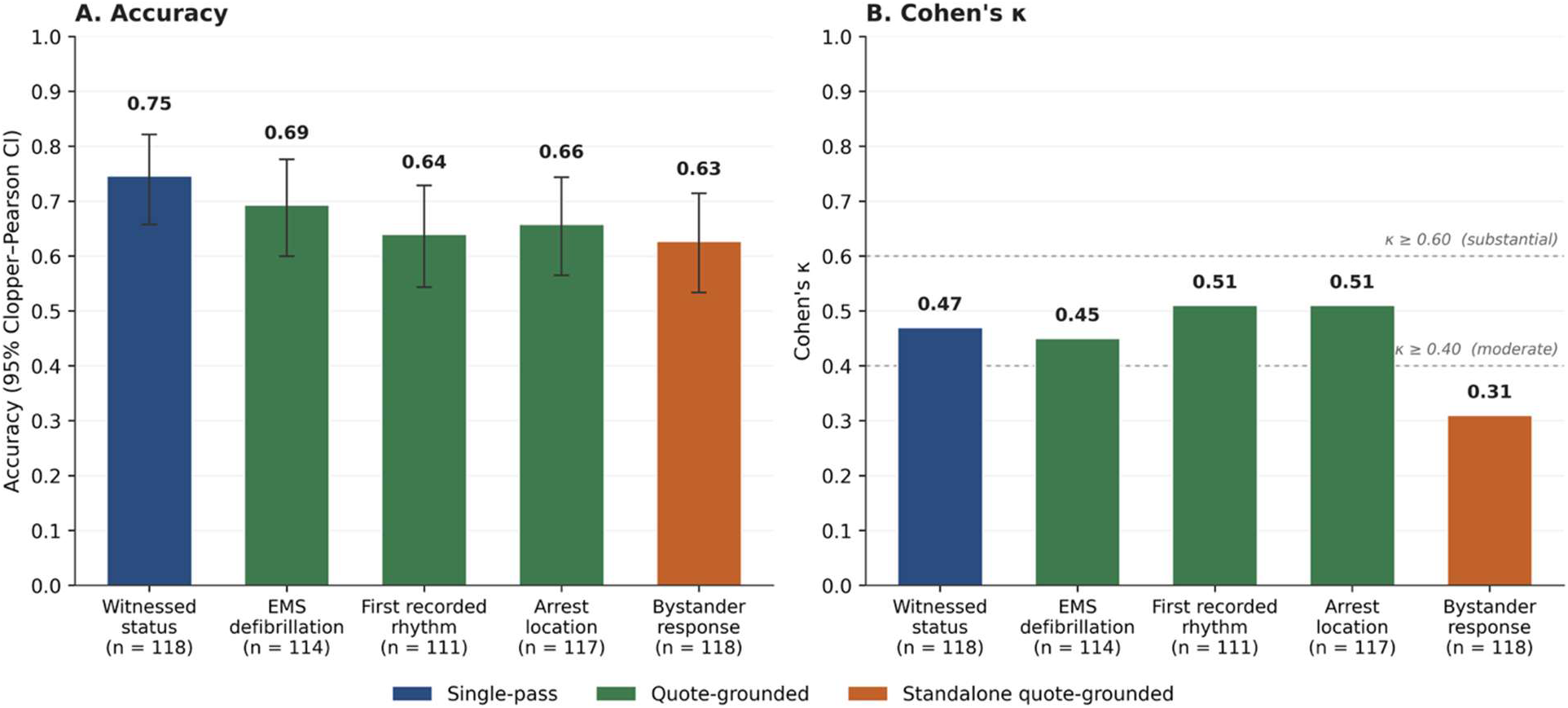
Variable-level performance. Figure 5 Legend: Variable-level accuracy (Panel A) and Cohen’s κ (Panel B) for the five extracted Utstein variables (witnessed status, EMS defibrillation, first recorded rhythm, arrest location, and bystander response) among confirmed OHCA encounters with both LLM and human-abstracted values present (n varies by variable; both-present analysis). Error bars denote 95% Clopper–Pearson exact binomial confidence intervals for accuracy. Bar color indicates extraction method (single-pass vs. quote-grounded vs. standalone quote-grounded). Landis–Koch thresholds for moderate agreement (κ ≥ 0.40) and substantial agreement (κ ≥ 0.60) are shown as dashed reference lines on Panel B. Abbreviations: OHCA, out-of-hospital cardiac arrest; LLM, large language model; EMS, emergency medical services; CPR, cardiopulmonary resuscitation; CI, confidence interval.

## Discussion

In this evaluation of OHCA-EXTRACT, an LLM-assisted pipeline identified true OHCA encounters from an ICD-flagged ED cohort with accuracy of 0.91 and specificity of 0.75 and extracted five core Utstein variables with accuracy ranging from 0.63 to 0.75. The performance of the identification module supports an LLM-based approach for potential development of administrative cohorts, while the Utstein variable extraction approach will require further methodological development and testing before it can substitute for human abstraction at scale. Prior LLM-based extraction efforts in oncology^11^, stroke^18^, and social determinants of health^13^ have demonstrated feasibility across clinical domains, but performance for OHCA workflows has not previously been rigorously evaluated, to our knowledge.^12,14^ The present work contributes a validated two-module LLM pipeline supporting future work to improve accuracy and further testing at scale. However, this work highlights both the promise and current limitations of LLM-assisted abstraction workflows. Several of our findings are worth discussing.

The OHCA identification module demonstrated high sensitivity (0.94) and moderate specificity (0.75). Within the development cohort, this performance profile is consistent with potential screening applications. However, because these metrics reflect iterative within-sample optimization rather than independent evaluation, generalization to external repositories requires prospective validation before screening utility can be established. Automated extraction of clinical variables from unstructured EHR documentation has previously been explored in clinical research, initially through rule-based NLP and domain-specific information-extraction systems, and more recently through supervised machine-learning classifiers and large language models.^19,20^ While rule-based NLP approaches offer interpretability and deterministic behavior, they require substantial customization and have limited capacity for temporal reasoning.^11,12^ Supervised classifiers trained on labeled clinical text improve on some of these limitations but are constrained by the availability and scale of annotated data and generalize poorly across institutions with differing documentation standards.^14^ A major barrier to observational OHCA research is reliable identification of cases. Because administrative codes lack specificity, there is a high risk for misclassification bias.

Although the identification module performed well in screening potential OHCA cases, performance of the variable-extraction module was more modest and highly dependent on the specific variable and extraction strategy. Agreement between the module and the human abstractors was moderate for most variables. Notably, witnessed status showed the highest accuracy under the single-pass approach but a lower kappa than the categorical variables. This pattern is consistent with binary variables with skewed prevalence. Whereas first recorded rhythm and arrest location achieved the highest chance-corrected agreement (κ = 0.51) using quote-grounded extraction. Constraining the model to provide explicit textual evidence for its outputs likely reduced the potential for hallucination and improved agreement for several variables. Even so, agreement remained low for bystander CPR (κ = 0.31), which may reflect ambiguity in clinical documentation and/or potential model error. One source of disagreement may be that the model is using only explicit textual evidence, whereas human abstractors may infer meaning from contextual clues.

Our model confidence scores were uniformly high, even when incorrect classifications were present. The confidence field in this pipeline reflects the model’s self-reported certainty within its structured JSON output and is not a calibrated probability estimate. With all scores at 0.9 or 1.0 and errors distributed uniformly across this narrow range, the confidence field functioned as a near-constant in this evaluation with no practical ability to discriminate correct from incorrect classifications. Importantly, the prespecified safety fallback was designed to trigger resubmission at confidence below 0.7. However this was never activated, confirming that confidence-threshold-based routing was unnecessary in the pipeline design.

Overall, our findings suggest a hybrid abstraction workflow, leveraging a human-in-the-loop approach and working in tandem with the LLM pipeline may yield the best results. LLMs may be able to screen large volumes of clinical data to identify potential OHCA cases of interest with greater specificity than conventional approaches such as ICD codes, while human abstractors may then focus on key variables of interest, especially those that may require interpretation of ambiguous documentation. The overall workload is reduced because the initial burden of case identification is shifted to the LLM pipeline, and human judgment is reserved for situations requiring critical and nuanced judgment. Because confidence scores demonstrated no discriminative value for error detection, human review cannot be triggered by confidence thresholds alone. Quality assurance in a deployed hybrid workflow should instead rely on pre-specified random output auditing, stratified review rates derived from variable-level error rates established in external validation, or documentation-completeness criteria as the basis for selective human verification. At the systems level, automation of OHCA data abstraction has significant and meaningful implications for quality improvement efforts and resuscitation science. Manual abstraction can be resource-intensive, limiting data collection in resource-constrained settings. By reducing the cost associated with data abstraction, LLMs may expand registry participation, reduce data missingness, and facilitate linkages between prehospital and in-hospital data. This pipeline is best positioned for sites and studies without dedicated OHCA abstractor resources. At sites with established registry infrastructure, prospective collection by trained personnel remains the reference standard for Utstein variable completeness and a tool such as OHCA-EXTRACT should be evaluated against that baseline before consideration of usage.

There are several areas for future work, including ways to improve performance and assess whether similar workflows may be applicable to other conditions. Improvements in upstream documentation (e.g., consistent documentation standards for both prehospital and in-hospital phases of care) or incorporation of more structured data elements may enhance performance of models. Further refinement of quote-grounded methods may address complex variables. Additionally, external validation across diverse cohorts and forms of documentation will be needed to ensure robustness prior to widespread clinical deployment. This will be important to address potential sources of bias both from the data and model side.

## Limitations

This study has several limitations. First, it was conducted within a single health system, and the generalizability of the pipeline to institutions with different documentation practices, patient populations, or EHR systems remains untested without external validation. Second, iterative refinement and final evaluation were conducted on the same cohort, with no separate development hold-out set. All reported metrics are resubstitution estimates that quantify performance on the data used to build the pipeline rather than on independent data. Such estimates are almost certainly optimistic relative to a prospectively collected or held-out cohort. Thus, these findings cannot be confirmed without further testing using an independent evaluation dataset. The performance figures reported here should also be interpreted as upper-bound estimates pending external validation and not as deployment-ready benchmarks. Third, the human-abstracted reference standard, while demonstrating substantial inter-abstractor agreement, retained variability among trained reviewers, particularly for fields with ambiguous documentation, indicating an upper limit on attainable agreement for any abstraction approach, automated or manual. Relatedly, we were unable to compute variable-specific inter-abstractor agreement for the five evaluated fields. The aggregate Fleiss’ κ (0.69) pools agreement is across all data-dictionary variables. We could therefore not establish the variable-level documentation ceiling against which the model’s per-variable agreement should be interpreted. This was particularly true for bystander CPR, where the low model agreement (κ = 0.31) may partly reflect ambiguous source documentation rather than extraction error alone. Fourth, several methodological constraints should temper interpretation. Only a single LLM (GPT-4o-mini) was evaluated, with no head-to-head comparison against larger frontier models. The non-OHCA subgroup was small (n = 28), yielding a wide specificity confidence interval (0.55–0.89) and clinical note inputs were truncated to 1,800 tokens. Although truncation is a plausible source of error in encounters with lengthy narratives, a focused audit of all false negatives found nonattributable to truncation. Each case the model’s rationale engaged directly with the documented arrest location, indicating the relevant text was retained within the input window. Fifth, model confidence scores showed a ceiling effect (all values 0.9–1.0), limiting any confidence-threshold–based human-review strategy. Because all model calls used non-deterministic infrastructure despite a temperature of 0, outputs are not bit-identical across executions. Repeat runs of the identification pipeline reclassified one to two of the 152 encounters, leaving accuracy unchanged (0.91) and shifting sensitivity, specificity, and predictive values by ≤ 0.04. Reported point estimates therefore carry minor run-to-run variability, while metric-level conclusions were stable across runs. Lastly, we evaluated only a subset of Utstein variables. LLM performance may differ for other fields, especially those requiring complex reasoning or synthesis across multiple documentation sources.

## Conclusion

An LLM-assisted pipeline identified true OHCA encounters within an ICD-flagged ED cohort with accuracy and predictive value exceeding an administrative-coding baseline, while automated extraction of five core Utstein variables achieved only fair-to-moderate agreement with human abstraction. These within-sample findings support further development of LLM-assisted, human-augmented abstraction workflows for OHCA surveillance, but external validation is required before generalization or deployment.

## Declarations

### Ethics approval and consent to participate

The study was reviewed and approved by the Institutional Review Board at the Icahn School of Medicine at Mount Sinai (IRB #STUDY-24-01201) and informed consent was not required.

### Declaration of generative AI and AI-assisted technologies

During the preparation of this work, the authors used a secure AI platform for formatting assistance of the final manuscript. After using this tool, the authors reviewed and edited the content as needed and take full responsibility for the content of the publication.

### Consent for publication

All authors consent to publication of this manuscript.

### Availability of data and materials

The data used for this study are not available for dissemination. All relevant code, figures, and methodological materials (large language model prompts and output schemas) are available at [repository link and DOI to be provided upon acceptance].

### Authors’ contributions

EEA and GN conceived and designed the study, developed the LLM pipeline architecture, led prompt engineering and schema design. DB and KP engineered the initial data extraction and pull from the electronic health record. EEA and ACS conducted data analysis and interpretation and drafted the manuscript. EEA led the overall study, secured funding, and supervised all phases of the work. GNN provided senior methodological and informatics mentorship and contributed to conception, design, and interpretation. DA, AS, KP, JJ, AZ, RH, BSA, and LDR contributed to interpretation of results, provided critical revisions to successive manuscript drafts, and approved the final version. All authors reviewed and approved the final manuscript.

## Acknowledgements

This work was supported in part through the computational and data resources and staff expertise provided by Scientific Computing and Data at the Icahn School of Medicine at Mount Sinai and supported by the Clinical and Translational Science Awards (CTSA) grant UL1TR004419 from the National Center for Advancing Translational Sciences. Research reported in this publication was also supported by the Office of Research Infrastructure of the National Institutes of Health under award numbers S10OD026880 and S10OD030463. The content is solely the responsibility of the authors and does not necessarily represent the official views of the National Institutes of Health.

## Abbreviations

OHCA: out-of-hospital cardiac arrest
LLM: large language model
EHR: electronic health record
EMS: emergency medical services
ED: emergency department
ICD: International Classification of Diseases
CPR: cardiopulmonary resuscitation
REDCap: Research Electronic Data Capture

